# Impact of Diagnostic Stewardship on Ordering Trends and Pathogen Yield from mNGS Studies

**DOI:** 10.1101/2024.04.19.24306038

**Authors:** Ryan C. Shean, Elizabeth Garrett, James Malleis, Joshua A. Lieberman, Benjamin T. Bradley

## Abstract

**Background:** Given the cost and unclear clinical impact of metagenomic next-generation sequencing (mNGS), laboratory stewardship may improve utilization. This study examines mNGS results from two academic medical centers employing different stewardship approaches.

**Methods:** 80 mNGS orders (54 CSF and 26 plasma) were identified from 2019 to 2021 at the University of Washington (UW), which requires director-level approval for mNGS orders, and the University of Utah (Utah), which does not restrict ordering. The impact of mNGS results and the relationship to traditional microbiology orders were retrospectively evaluated.

**Results:** 19% (10/54) CSF and 65% (17/26) plasma studies detected at least one organism. Compared to CSF results, plasma results were more frequently clinically significant (23% vs 7%) and led to more novel diagnoses (15% vs 0%). Results affecting antibiotic management were more common for plasma than CSF (32% vs. 2%). Stewardship practices were not associated with statistically significant differences in results or antimicrobial management. The number and cost of traditional microbiology tests at UW was greater than Utah for CSF mNGS testing (UW: 46 tests, $6237; Utah: 26 tests, $2812; p<0.05) but similar for plasma mNGS (UW: 31 tests, $3975; Utah: 21 tests, $2715; p=0.14). mNGS testing accounted for 30-50% of the total microbiology costs.

**Conclusions:** Improving the diagnostic performance of mNGS by stewardship remains challenging due to low positivity rates and difficulties assessing clinical impact. From a fiscal perspective, stewardship efforts should focus on reducing testing in low-yield populations given the high costs of mNGS relative to overall microbiology testing expenditures.

## Introduction

Next-generation sequencing (NGS) is a powerful technology that allows for millions of DNA fragments to be independently and simultaneously sequenced. Metagenomic NGS (mNGS) is the analytical process by which all nucleic acid can be sequenced and classified, permitting complex populations of nucleic acid from many organisms to be analyzed in a single sample. These technologies have emerged as exciting, but expensive tools to diagnose infections from diverse anatomic sites and specimen types, including cerebrospinal fluid (CSF) and plasma.^1–6^ An advantage of mNGS over traditional microbiological approaches is the ability to detect any bacterial, viral, and fungal organism, without explicit inclusion in the provider’s differential diagnosis.^3,7^ This stands in contrast to traditional microbiology testing, including culture, serology, and nucleic acid amplification tests, which depends on the provider’s differential diagnosis to help select the appropriate tests^8,9^. Plasma mNGS can also provide a non-invasive alternative to biopsy for the identification of deep infections such as invasive fungal disease or osteomyelitis and occult infections such as culture-negative endocarditis.^10–12^

However, the role of mNGS technology in infectious disease diagnostics and its place in the context of traditional microbiology testing is still unclear. As with other approaches, mNGS can identify clinically irrelevant targets such as colonizing or transient organisms. DNA can also persist well after an organism has been cleared; therefore, mNGS results must be interpreted in the context of the patient’s clinical presentation.^6^ Recent studies have described variable clinical utility of mNGS results, finding 12-55% of positive results to be clinically actionable^13,14^. This variable clinical impact is due to the diverse range of patient populations, clinical syndromes, and matrices that can be assessed with this unbiased diagnostic approach.^10,14–16^ Interpretation of the results is further complicated by differences in the analytic techniques and databases used to classify nucleic acid sequences^17^.

Due to the novelty, cost, limited clinical impact, and difficulty in properly interpreting mNGS results, a variety of stewardship practices have been implemented to improve test utilization^18^. Historically, diagnostic stewardship has been used in clinical microbiology testing to counterbalance well-intentioned but misdirected use^19^. Some successful implementations of diagnostic stewardship include decision support testing (e.g., hard stops on *Clostridioides difficile* testing on formed stool samples) as well as automated reflex testing to ensure that proper algorithms are correctly followed^20^. There is a clear need for evidence-based guidance to help inform when mNGS infectious disease testing will improve patient outcomes^19^. Currently available data show diagnostic utility largely depends upon the use case in which mNGS is performed^14,21,22^. To properly inform diagnostic stewardship guidelines, data such as the cost, timing, and context of mNGS ordering relative to traditional microbiology testing is needed.

In this retrospective analysis, we examined CSF and plasma mNGS orders in patients across two different multicenter hospital systems, which provide extensive specialty care for cancer, transplant, and trauma patients. We evaluated mNGS assay performance, impact on antibiotic management, and relationship to traditional microbiology testing in terms of timing and cost. We highlight the impact that stewardship may have on these measures to aid laboratories in the establishment of evidence-based guidelines.

## Materials and Methods

### Ethics Statement

This study was approved by the University of Washington (UW) Institutional Review Board Committee (IRB ID STUDY00010777) and University of Utah (Utah) Institutional Review Board (IRB 00151187) as waived research; informed consent was not required.

### mNGS Testing

CSF mNGS was performed by the University of California San Francisco Clinical Microbiology Lab (California); plasma mNGS was performed by Karius (Redwood City, California). Both laboratories are Clinical Laboratory Improvement Amendments (CLIA)-certified. Samples were submitted in accordance with the specimen requirements for each laboratory.

### Institutional Approval Processes for mNGS

At UW, the interdisciplinary Laboratory Formulary Committee sets an annual limit on the number of mNGS send-out tests and instituted an approval process. Briefly, provider requests for CSF and plasma mNGS testing underwent a collaborative review process between clinical microbiology directors and the care team with required input from infectious disease physicians. Considerations for approval included: 1) expected impact to clinical decision making, 2) traditional microbiologic testing results (cultures, serology, and nucleic acid amplification test (NAAT)), 3) clinical suspicion for infectious etiology, and 4) difficulty of obtaining lesional tissue. Extended factors included underlying medical conditions, immunocompromised status, and recent antibiotic exposure. At Utah, laboratory approval for CSF and plasma mNGS was not required prior to test ordering.

### Data Collection and Analysis

Specimens approved for CSF or plasma mNGS testing at UW from July 2019 to March 2021 and ordered at Utah from January 2019 to December 2021 were retrospectively identified through the laboratory information systems (LIS). Patient charts were reviewed to determine the significance and impact of mNGS testing. Any test result in which one or more organisms were detected by the assay were considered a positive mNGS result. Clinically significant results were adjudicated based on review of the medical chart and impression of the clinical team caring for the patient. Organisms known to cause latent infections (e.g. CMV, EBV) or part of the oral or gastrointestinal flora (e.g., coagulase-negative staphylococci, viridians streptococci) were considered clinically insignificant unless otherwise indicated by the medical team or supported by orthogonal clinical testing. Novel pathogens were defined as organisms deemed clinically significant and not observed by orthogonal means prior to mNGS results. Clinical impact was evaluated based on as alterations in antimicrobial treatment (escalation, de-escalation, or change) that specifically considered mNGS results, positive or negative. Additional data were gathered from the LIS regarding patient demographics and traditional microbiology orders placed. For this study’s purposes, traditional microbiology orders were considered any test (culture, serology, antigen, or molecular assay) ordered to assist in the clinical diagnosis excluding mNGS. Surveillance and routine screening studies were excluded. Testing turn-around time (TAT) was calculated from the time the specimen was accessioned to the time the result was received from the performing lab.

The billing charges associated with infectious disease testing for patients in this study were evaluated using the hospital fee schedule. In order to standardize cost estimates across both health systems, the UW hospital chargemaster from November 2020 was used for both UW and Utah patients^23^. For Utah patients who received testing not available on the UW chargemaster, estimated cost of microbiology testing was obtained using the ARUP cost estimator tool (https://www.aruplab.com/testing/resources/calculator). Costs between the UW chargemaster and the ARUP cost estimator tool were generally very similar for tests offered by both institutions which also matched closely with the Medicare reimbursement schedule. Only microbiological diagnostic costs from the hospitalization during which mNGS was ordered were included in the analysis, except for one patient who had two hospitalizations 9 days apart for the same clinical syndrome and has tests from both stays included.

Data was analyzed in R (v4.2). Statistical tests performed included Pearson’s Chi-squared test with Yates’ continuity correction and Welch’s two sample t-tests. Statistical significance was set at a threshold of p <0.05.

## Results

### Patient Demographics and mNGS Results

During the study period, a total of 80 mNGS tests met inclusion criteria, 28 from the UW hospital system and 52 from the Utah hospital system (Table 1). For the UW cohort, mNGS testing was rejected in 12 instances. Reasons included alternative testing methods available in-house (n=5), low concern for an infectious agent (n=3), and a suspected pathogen already identified (n=2). Male and female individuals were equally represented in both CSF mNGS cohorts; however, a higher proportion of males received plasma mNGS studies (81% male vs 19% female). The plasma mNGS cohort included a greater number of immunocompromised individuals relative to the CSF mNGS cohort (77% vs 39%). Excluding the Utah CSF mNGS group, antibiotics were administered to over half of the individuals in each cohort prior to mNGS specimen collection and in 70% of those with a positive mNGS result. At least one individual in each cohort received mNGS testing as an outpatient.

**Table 1.** Demographic and Clinical Information for Individuals Receiving mNGS.

|  | CSF |  |  | Plasma |  |  |
| --- | --- | --- | --- | --- | --- | --- |
|  | Utah | UW | Total | Utah | UW | Total |
| Individuals | 44 | 10 | 54 | 8 | 18 | 26 |
| Median Age (IQR) | 45 (33-46) | 47 (34-51) | 45 (35-64) | 52 (41-49) | 52 (41-57) | 53 (41-62) |
| **Sex (n, [%])** |  |  |  |  |  |  |
| Male | 18 (41%) | 4 (40%) | 22 (41%) | 8 (100%) | 13 (72%) | 21 (81%) |
| Female | 26 (59%) | 6 (60%) | 32 (59%) | 0 (0%) | 5 (28%) | 5 (19%) |
| **Clinical Information (n, [%])** |  |  |  |  |  |  |
| Immunosuppressed | 15 (34%) | 4 (40%) | 21 (39%) | 7 (88%) | 13 (72%) | 20 (77%) |
| Received antibiotics | 12 (27%) | 8 (80%) | 20 (37%) | 5 (63%) | 17 (94%) | 22 (85%) |
| Outpatient | 7 (16%) | 1 (10%) | 8 (15%) | 1 (13%) | 2 (11%) | 3 (12%) |
| **Clinical Indication (n, [%])** |  |  |  |  |  |  |
| Neurologic | 44 (100%) | 10 (100%) | 54 (100%) | 4 (50%) | 1 (6%) | 5 (19%) |
| Endovascular | 0 (0%) | 0 (0%) | 0 (0%) | 2 (25%) | 11 (61%) | 13 (50%) |
| Pulmonary | 0 (0%) | 0 (0%) | 0 (0%) | 1 (13%) | 3 (17%) | 4 (15%) |
| FUO | 0 (0%) | 0 (0%) | 0 (0%) | 1 (13%) | 2 (11%) | 3 (12%) |
| **Results (n, [%])** |  |  |  |  |  |  |
| Positive mNGS results | 10 (23%) | 0 (0%) | 10 (19%) | 6 (75%) | 11 (61%) | 17 (65%) |
| --Clinically significant | 4 (9%) | 0 (0%) | 4 (7%) | 1 (13%) | 5 (28%) | 6 (23%) |
| --Novel | 0 (0%) | 0 (0%) | 0 (0%) | 0 (0%) | 4 (22%) | 4 (15%) |
| Change in antibiotic management | 1 (2%) | 0 (0%) | 1 (2%) | 2 (25%) | 6 (33%) | 8 (31%) |

Clinical indications for CSF mNGS testing were limited to individuals presenting with neurologic conditions which included clinical features of meningitis or encephalitis, radiologically evident lesions or vasculitis, and sub-acute cognitive impairment. Plasma mNGS was ordered for a wider range of indications including endovascular disease such as culture negative endocarditis (50%, 13/26), neurologic dysfunction (19%, 5/26), pulmonary lesions with negative microbiologic culture or contraindications for lesion biopsy (15%, 4/26), fever of unknown origin (12%, 3/26), and liver abscess (4%, 1/26).

At least one organism was detected in 19% (10/54) of CSF specimens and 65% (17/26) of plasma specimens. Overall, plasma mNGS results were more likely to identify clinically significant pathogens compared to CSF (23% vs 7%). Plasma mNGS was also more likely to identify novel pathogens compared to CSF mNGS (15% vs 0%). While differences between study site results did not reach statistical significance, clinically significant organisms from plasma were observed at twice the frequency in the UW cohort (28%, n= 5) as compared to the Utah cohort (13%, n= 1). Similarly, all novel mNGS results (n=4) occurred in the UW plasma CSF group. Prior antibiotic use was not associated with a decreased likelihood of detecting a clinically significant pathogen.

Alterations in antimicrobial therapy were infrequent within the CSF mNGS cohort with only one result leading to a change in therapy. Antibiotic management changes were more frequent in the plasma mNGS cohort with 31% (8/26) of results leading to a change in management. Plasma mNGS results also demonstrated a similar impact on management between Utah and UW (25% and 33%). In 44% (4/9) of cases where mNGS results affected antibiotic management, the change was made following a negative result (n=1) or identification of a non-significant pathogen (n=3) (Supplemental Table 1).

### Ordering Practices and Costs for Conventional Microbiology Testing

A common criticism of mNGS is the high cost of a single test. The average cost of plasma mNGS and CSF mNGS in this study was $2000 and $2900, respectively. We compared the volume and associated costs of other microbiology testing ordered for the same clinical syndrome which led to use of mNGS (Figure 1). A significant difference was seen between study sites for CSF mNGS testing (UW: 46 tests, $6237; Utah: 26 tests, $2812; p<0.05) but not for plasma mNGS testing (UW: 31 tests, $3975; Utah: 21 tests, $2715; p=0.14) (Table 2, Figure 2A). The greater number of total tests in the UW CSF mNGS group was due to a comparatively higher number of tests ordered prior to mNGS testing (UW: 40; Utah: 20, p<0.05). In the period following CSF mNGS, testing volumes were similar between sites (UW: 6; Utah: 6, p=0.89). In relative terms, mNGS accounted for 30-50% of the total microbiology testing costs regardless of test or stewardship practice (Figure 2B). While CSF mNGS was ordered earlier for Utah patients than for UW patients (4.5 vs 14 days), the majority were discharged by the time results were available whereas UW patients were more likely to remain inpatients (52% vs 10%). For plasma mNGS, 0% (0/8) of Utah patients and 6% (1/18) of UW patients were discharged at time of mNGS result.

**Table 2.** Traditional Microbiology Test Volume and Cost for Individuals Receiving mNGS.

|  | CSF |  |  | Plasma |  |  |
| --- | --- | --- | --- | --- | --- | --- |
|  | Utah | UW | p-value | Utah | UW | p-value |
| **Micro tests ordered (mean, [SD])** |  |  |  |  |  |  |
| Total | 26 (18) | 46 (18) | **0.01** | 21 (8) | 31 (25) | 0.14 |
| Pre-mNGS | 20 (12) | 40 (15) | **0.005** | 16 (8) | 23 (13) | 0.15 |
| Post-mNGS | 6 (12) | 6 (4) | 0.89 | 5 (6) | 9 (19) | 0.43 |
| **Cost (mean, [SD])** |  |  |  |  |  |  |
| Total | 2812 (2007) | 6237 (2923) | **0.008** | 2715 (1223) | 3975 (524) | 0.37 |
| Pre-mNGS | 2147 (1392) | 5123 (2160) | **0.003** | 2125 (1096) | 2399 (1965) | 0.67 |
| Post-mNGS | 665 (1171) | 811 (878) | 0.67 | 589 (804) | 1575 (4552) | 0.41 |
| **mNGS ordering** |  |  |  |  |  |  |
| Hospital Day mNGS ordered (range) | 4.5 (0-21) | 14 (2-80) | **0.03** | 12 (7-24) | 6.5 (2-24) | 0.38 |
| Median TAT, days (range) | 14 (9-26) | 14 (6-21) | 0.65 | 1 (1-4) | 3 (2-7) | **0.05** |
| Discharged at time of result, n | 23 | 1 | **0.03** | 0 | 1 | 0.79 |

**Figure 1.**
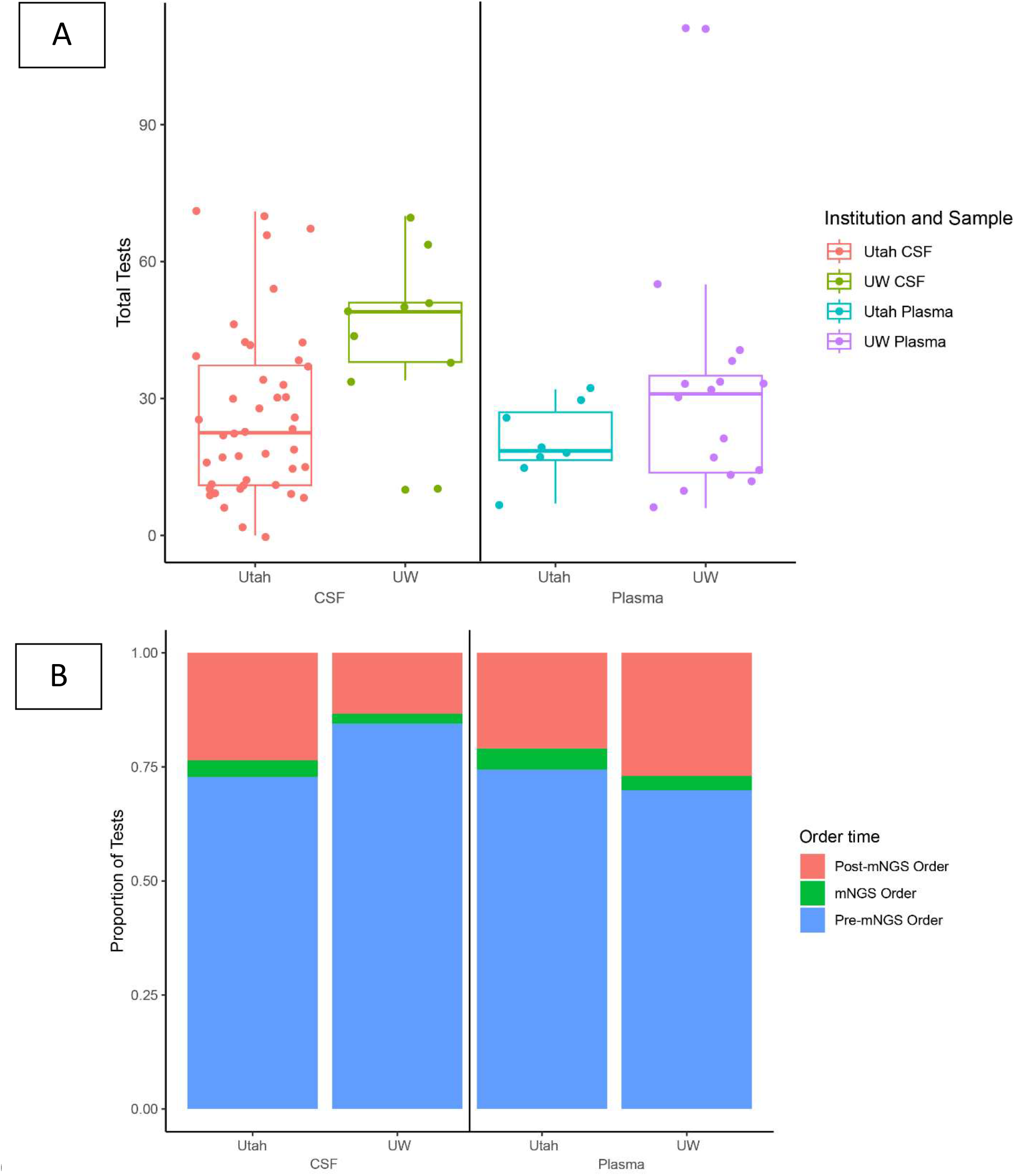
Total and Relative Volumes of Traditional Microbiology Test Orders for Patients Receiving mNGS A) Traditional microbiology testing volumes for patients undergoing CSF or plasma mNGS studies. Results separated by study site and test. Box and whisker plot represents mean, IQR, and 95^th^ percentile. B) Average proportion of total traditional microbiology tests ordered before and after plasma or CSF mNGS at Utah and UW. The mNGS order represents the relative size of a single test order.

**Figure 2.**
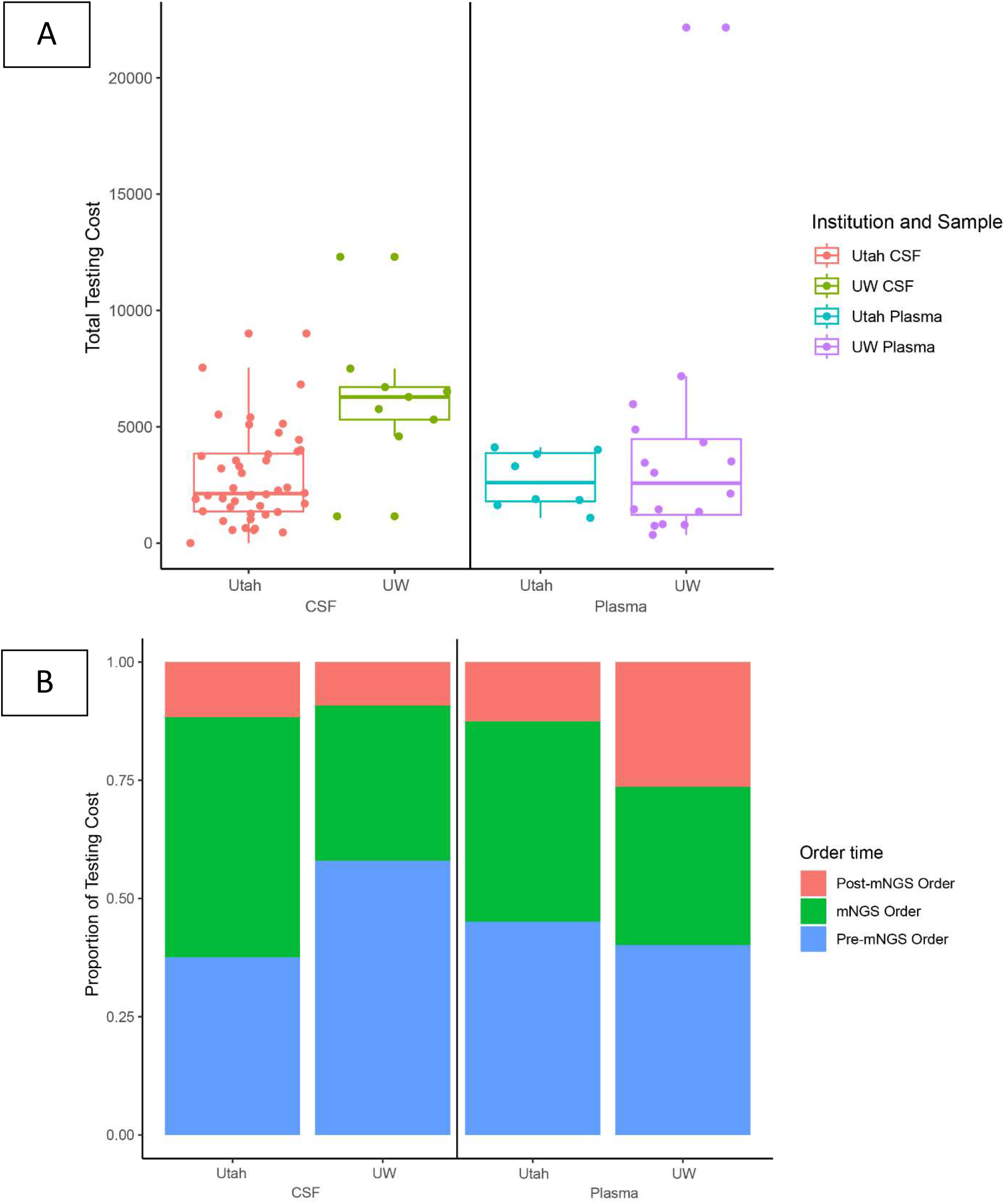
Total and Relative Cost of Traditional Microbiology Test Orders for Patients Receiving mNGS A) Traditional microbiology testing costs for patients undergoing CSF or plasma mNGS studies. Results separated by study site and test. Box and whisker plot represents mean, IQR, and 95^th^ percentile. B) Cost of traditional and mNGS orders relative to the overall microbiology testing expenditure.

Traditional microbiology orders varied relative to the type of mNGS test (CSF vs. plasma) and timing (pre vs. post-mNGS) (Table 3). For individuals undergoing CSF mNGS, the two most common traditional microbiology tests were CSF culture and blood culture. Tests that were relatively more common after CSF mNGS ordering included tissue bacterial cultures and tissue fungal cultures. For plasma mNGS orders, the most common tests overall were bacterial and fungal cultures from blood. As was observed in the CSF mNGS cohort, tests from invasively collected specimens such as tissue bacterial cultures and tissue fungal cultures were ordered more frequently following plasma mNGS orders.

**Table 3.** Most Common Traditional Microbiology Tests Ordered for Individuals Receiving mNGS.

| CSF |  |  |  |  |  |
| --- | --- | --- | --- | --- | --- |
| Pre-mNGS |  | Post-mNGS |  | Total |  |
| Test | Number | Test | Number | Test | Number |
| CSF culture | 81 | AFB culture | 17 | CSF culture | 92 |
| Blood culture | 64 | Blood culture | 17 | Blood culture | 81 |
| HSV PCR | 55 | Tissue culture | 15 | AFB culture with stain | 68 |
| AFB culture | 51 | CSF culture | 11 | HSV PCR | 63 |
| VZV EIA | 48 | CMV PCR | 10 | Cryptococcal Antigen | 53 |
| Cryptococcal Antigen | 47 | Tissue fungal culture | 10 | VZV EIA | 51 |
| CSF multiplex PCR | 45 | Bacterial culture, body fluid | 9 | CSF multiplex PCR | 48 |
| CSF fungal culture | 40 | CSF culture, anaerobic | 8 | CMV PCR | 44 |
| VZV PCR | 38 | HSV PCR | 8 | CSF fungal culture | 44 |
| CMV PCR | 34 | Coccidioides Ab compfix | 8 | VZV PCR | 44 |
| Plasma |  |  |  |  |  |
| Pre-mNGS |  | Post-mNGS |  | Total |  |
| Test | Number | Test | Number | Test | Number |
| Blood culture | 128 | Blood culture | 20 | Blood culture | 148 |
| Blood fungal culture | 27 | Broad-range fungal PCR | 15 | Blood fungal culture | 28 |
| AFB culture | 13 | M. tb complex PCR | 14 | AFB culture | 25 |
| Aspergillus galactomanin EIA | 12 | Broad-range bacterial PCR | 14 | Broad-range fungal PCR | 22 |
| Urine Culture | 11 | AFB culture | 12 | Broad-range bacterial PCR | 20 |
| Brucella Ab screen, agglutination | 9 | Tissue culture | 12 | Tissue culture | 19 |
| Bartonella Ab panel | 9 | NTM PCR | 12 | M. tb complex PCR | 16 |
| Q fever Ab, IgG and IgM | 9 | Tissue fungal with direct exam | 6 | NTM PCR | 14 |
| EBV quant PCR | 8 | Wound culture, anaerobic | 4 | Urine Culture | 12 |
| Nocardia Culture | 7 | Brucella Ab screen, agglutination | 3 | Aspergillus galactomanan EIA | 12 |

## Discussion

The introduction of mNGS into clinical microbiology represents an exciting prospect for the diagnosis of infectious disease. However, the role of this technology remains a matter of discussion, as is reflected by the variable performance of mNGS in clinical practice.^21,24^ In this study we found that 19% (10/54) of CSF results and 65% (17/26) or plasma results were positive but only 2% (1/54) of CSF results and 31% (8/26) of plasma results led to a change in antimicrobial therapy. Stewardship was not associated with a statistically higher rate of results which changed management. These results are similar to other studies which have demonstrated a low return of clinically actionable results from mNGS.^10,12,14^

While improving diagnostic yields may prove challenging, a more attainable goal for stewardship programs can be the reduction of low-yield testing. The advantages of stewardship are best highlighted when comparing CSF mNGS patient populations between Utah and UW. At Utah, where ordering was not regulated, 68% (30/44) of patients were either tested as outpatients or discharged by the time results were available. This stands in contrast to UW where only 20% (2/10) met these criteria. It could be argued that one potential benefit of the policy at Utah was that mNGS was performed earlier (4 vs. 14 days) and with fewer traditional microbiology studies (26 vs. 46) relative to UW, thus increasing the likelihood of yielding novel or actionable results. However, within the Utah cohort no novel diagnoses were made and antibiotic management was only affected in one case following a negative result.

The CSF mNGS test was developed to detect esoteric and infrequent pathogens in patients with a high likelihood of unexplained infectious meningitis/encephalitis. However, in a significant number of cases the assay was used as part of a ‘rule-out’ approach prior to initiation of biologics for autoimmune encephalitis. Limiting these orders is critically important as data suggest that CSF mNGS may not achieve the sensitivity sufficient to rule-out infection as compared to traditional microbiological diagnostics.^1,14^ In our data, we also observed a high rate of non-clinically significant organisms identified from plasma mNGS. Reducing the risk of potential false positive and false negative results is best achieved by limiting mNGS studies to populations with a high pre-test probability of infection.^12,14^

Interestingly, we observed a significantly longer TAT for plasma mNGS at UW as compared to Utah. We hypothesize that specimens were collected and then held while the expert committee reviewed the case and/or awaited results from pending, conventional microbiology tests. If correct, the additional time due to stewardship activities may account for the longer TAT at UW.

This study provides a starting point for analyzing the impact of stewardship on the overall cost of microbiology testing in patients receiving mNGS. While the total microbiology testing costs were lower per individual in the unsupervised setting at Utah, this fails to account for cost saving generated by avoiding unnecessary mNGS studies in low-yield patients. For this reason, costs saving should be assessed at the systems level and not the patient level. Another important finding of our study is that the cost of a single mNGS assay is similar to the total cost of all other microbiology tests ordered.

Therefore, requiring more traditional microbiology testing prior to mNGS ordering is likely a cost-effective policy. However, this approach risks potential delays in diagnosis if negative cultures or finalized results are required prior to mNGS approval. Specific clinical use cases for mNGS and pre-requisite traditional microbiology studies should be established in a multidisciplinary process prior to testing.

Several limitations exist in this study; first is the retrospective nature of the data collected and the post-hoc assessment of mNGS results on antibiotic management. We attempted to use a narrow, but well-defined criteria that could be supported by the clinical record. However, mNGS results may be assessed for much wider clinical impacts and the impact of a negative result may be difficult to determine from the clinical record.^14^ Specifically, assessing the clinical impact of a negative CSF result from a patient with a pre-existing low probability of infection remains challenging and will require studies constructed to examine this question. While this study provides cost estimates from two different hospital systems, it is possible these estimates may not be widely generalizable between hospital systems. Individual nuances including clinical service leadership and physician ordering practices hinder direct comparisons to this study. It is important for any system considering mNGS stewardship to realize policies will need to be built on an individual basis and tailored to the patient population.

This study provides several lessons for developing mNGS stewardship policies. First, clinical performance may not be the primary endpoint to evaluate the success of mNGS stewardship as the likelihood of positive results or changes in management are low regardless of approach. Instead, programs should focus on reducing testing in low-value situations, specifically for patients who will be discharged prior to the results becoming available and in patients for whom “rule-out” testing is requested. This latter situation was commonly observed when patients underwent CSF mNGS studies prior to initiation of immunosuppressants. Examining the financial impacts of mNGS stewardship should be examined in aggregate, and costs savings associated with preventing low-yield mNGS testing should be noted. The threshold for suggesting additional pathogen-directed testing prior to mNGS should be low given mNGS may account for 30-50% of total testing costs while marginally improving detection of novel pathogens. Finally, decisions for mNGS send-out testing approval should be made expeditiously to prevent testing delays.

## Data Availability

All data produced in the present study are available upon reasonable request to the authors

